# Immunochemotherapy as induction treatment in Stage III (N2, N3) Non-small cell lung cancer

**DOI:** 10.1101/2021.06.03.21257757

**Authors:** Hongsheng Deng, Hengrui Liang, Wei Wang, Jianfu Li, Shan Xiong, Bo Cheng, Caichen Li, Qing Ai, Zhuxing Chen, Haixuan Wang, Wenhua Liang, Jianxing He

## Abstract

**Background:** To increase locoregional and systemic tumor control, a portion of patients with stage III (N2, N3) non-small cell lung cancer (NSCLC) received pulmonary resection after immunochemotherapy in our center. Herein, we assessed the real-world downstage (T, N stage) effectiveness of immunochemotherapy as induction treatment and explored the proper cycle number for stage III (N2, N3) NSCLC.

**Methods:** Biopsy confirmed stage III (N2, N3) NSCLC patients who underwent immunochemotherapy between January 1^st,^ 2018, to August 30^th,^ 2019, were identified. Tumor radiologic regression, lymph node down-staging, and pathological response information were collected.

**Results:** In total, 16 patients with stage IIIA NSCLC, 30 with stage IIIB NSCLC, 9 with stage IIIC NSCLC (N2, N3 metastasis) were included. After immunochemotherapy, 25/55 (45.5%) patients achieved an objective response. Ultimately, 33/55 (60.0%) patients received lobectomy plus systemic lymphadenectomy, of whom 18/33 (54.5%) obtained major pathological response (MPR) of the primary lesion, and 24 (72.7%) had pathological-confirmed lymph node downstage (N2-3 to N0-1). Notably, four patients had MPR of the primary lesion but without lymph node downstage. At the time of data cutoff (December 30^th,^ 2020), the median follow-up duration was 9.2 months (IQR 8.0-11.7), 24/33 (72.7%) of patients that had pulmonary resection were progression-free, with 30 of them alive. Binary logistics analysis showed that 3-4 induction cycles were favorably associated with MPR than 1-2 cycles (p = 0.017).

**Conclusions:** Immunochemotherapy as induction treatment showed encouraging MPR and lymph nodes down-staging rates in stage III (N2, N3) NSCLC in this study. Prolonged (3-4) cycles of immunochemotherapy were recommended for a better pathological response.

## Introduction

Non-small-cell lung cancer (NSCLC) accounts for 80–85% of all lung cancer cases. Approximately 20% of patients with NSCLC are locally advanced disease at diagnosis and are generally considered inoperable. Outcomes remain poor for this subset of patients, with a median progression-free survival of 13 months and 3-year overall survival of 30%. Most locally advanced NSCLC experience disease progression, despite definitive concurrent chemoradiotherapy [1].

Anti-PD-1 immunotherapies (IO), which block the binding of the PD-1 receptor and its ligands (PD-L1/2), have been proven to improve the outcomes of patients with advanced NSCLC. Recently, the results of some phase II studies investigating the role of immunotherapy plus chemotherapy[2–4] or dual checkpoint inhibition [5] have been published, which supported the addition of IO in neoadjuvant treatment for patients with resectable stage NSCLC. The NADIM trial [6] assessed the efficacy and safety of neoadjuvant immunochemotherapy followed by surgery and adjuvant nivolumab in stage IIIA NSCLC patients. Down-staging occurred in 90% of cases, and 35 (85.4%) of 41 patients survived without recurrence following surgery. The phase II NEOSTAR trial [5] administrated three doses of nivolumab with or without ipilimumab as a neoadjuvant regiment in 44 patients with stage I–IIIA NSCLC. MPR rates in the nivolumab and nivolumab plus ipilimumab groups were 17% and 33%, respectively. However, advanced stage III NSCLCs with N2, N3 metastasis were generally excluded from trials respecting neoadjuvant immunochemotherapy due to the initial unresectability.

Long-term outcomes from phase III clinical trials of PD-1 inhibitors in previously treated patients with advanced NSCLC demonstrated a 2-year overall survival of 23%-29% and 5-year overall survival of 16%[7, 8]. A subsequent study, the phase III clinical trials PACIFIC, have released its latest outcome that 3-year overall survival of the durvalumab group attained 57.0% versus 43.5% of placebo[9]. The survival advantage with anti-PD-1 agents after concurrent chemoradiotherapy in patients with stage III, unresectable NSCLC has been proven. This pattern, which is able to cause long-term tumor regression and potential cure for advanced NSCLC, may render inoperable NSCLC operable. Chaft et al[10] reported five patients with metastatic cancer that underwent tumor resection following ICIs therapy. Surgery was carried out successfully, and 2 (40%) patients had pCR, with four patients remained disease-free 7-23 months postoperatively. However, so far, no study with a sufficient sample size has demonstrated the down-staging (T, N stage) rate in stage III (N2, N3) NSCLC treated by induction immunochemotherapy due to low surgery rate in this cohort. And the long-term survival benefit of surgery following immunochemotherapy in stage III (N2, N3) NSCLC remains poorly elucidated.

Furthermore, since the proper cycle number of PD-1 blockades could not be explored from randomized trials, whether prolonged cycles of immunochemotherapy had better efficacy for tumor downstage remains unclear. Immunochemotherapy was administrated in initial stage III NSCLC patients with a potentially curable possibility in our center [11]. And with the aims of increasing both locoregional and systemic control, a portion of patients received radical tumor resection after immunochemotherapy. This retrospective study aimed to assess the real-world downstaging (T, N stage) effectiveness of induction immunochemotherapy for stage III NSCLC with N2, N3 metastatic; figure out the relationship between prolonged cycle and pathological response; with particular attention given to progression-free survival within a year of following up.

## Materials and Methods

### Study design and patient inclusion

Flow chart of the study design, inclusion, and exclusion criteria used for screening patients, and main oncological outcomes of included patients was shown in Figure 1. The study protocol and methods were reviewed by the institutional ethics committee of the First Affiliated Hospital of Guangzhou Medical University. Patients who underwent at least two cycles of PD-1 immuno-chemotherapy (PD-1 blockades included: Pembrolizumab, Nivolumab, Toripalimab, Camrelizumab or Sintilimab) plus platinum-based chemotherapy to decrease tumor size and down-staging lymph node (intention to surgery) at The First Affiliated Hospital of Guangzhou Medical University between January 1^st,^ 2018 to August 30^th,^ 2019 were identified and included. The data of this patient cohort consecutively retrospectively collected through electronic medical records.

**Figure 1.**
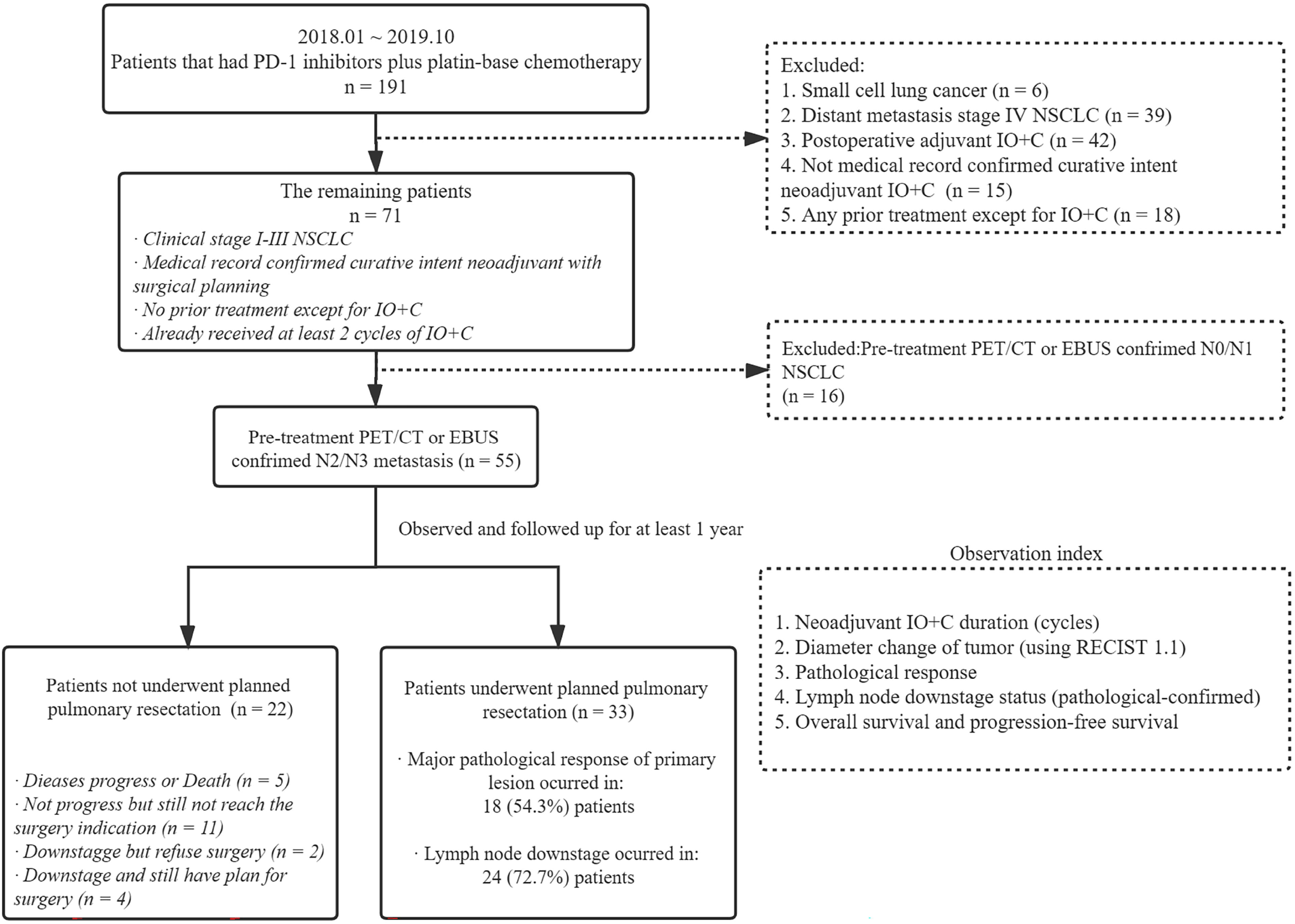
Flow chart of the study design, inclusion and exclusion criteria used for screening patients, and main oncological outcomes of included patients.

#### Exclusion criteria

patients with small-cell lung cancer (SCLC); patients diagnosed with stage IV NSCLC; or previously treated patients.

All patients were followed up until December 30^th,^ 2020.

### Immunochemotherapy and surgical technique

Specimens for cytological and histological examination were obtained via bronchoscopy before induction immune-chemotherapy. PD-L1 expression test was not compulsive in every case. Platinum-based doublet chemotherapy was prescribed every 21 days [12] with PD-1 inhibitors (Pembrolizumab, Nivolumab, Toripalimab, Tislelizumab, Camrelizumab, or Sintilimab). All patients underwent standard diagnostic and staging procedures. Computed tomography (CT) of the brain, chest, and abdomen were performed to exclude distant metastasis. CT scan was administrated every two cycles and at the last planned cycle of immune-chemotherapy. Lymph node status was assessed via PET-CT to first- and re-stage patients. All patients were confirmed to have no targetable driven mutations such as EGFR, ALK, ROS1, and BRAF.

More cycles were offered until the tumor was down-staged and became resectable. The resectability of patients was discussed by the multidisciplinary tumor board, which contained an expert group of thoracic surgeons, oncologists, and radiologists. Candidates for operation after induction immunochemotherapy: (1) Confirmed lymph node down-staging (assessed by PET/CT), the operation time will be at the second month after the last cycle of induction immune-chemotherapy; (2) complete cardiovascular examination tests, namely cardiopulmonary exercise test, echocardiogram and coronary angiography, pulmonary function tests showed tolerable cardio-pulmonary function for surgery.

All surgeries were initially attempted under video-assisted thoracoscopic surgery (VATS), converting to hybrid VATS or open surgery when necessary. Wedge resection was first performed; residual disease was excluded by frozen section evaluation. Then a lobectomy combined with systematic hilar and mediastinal lymphadenectomy with dissection of stations 2, 4, 7, 8, 9, and 10 during a right pneumonectomy and of stations 4, 5, 6, 7, 8, 9, and 10 during a left pneumonectomy. Double sleeve resection was performed in patients that did not tolerate pneumonectomy, and if the main bronchus infiltration by the tumor at the level of origin of the upper lobar bronchi exceeds 2 cm or involvement of pulmonary artery exceeds 3 cm, lung auto-transplantation would be the choice. The anastomosis was covered by the interposition of the vascular pedicled thymic flap, prepericardial fat, thymus, or mediastinal pleura in selected cases.

After the operation, the patients were provided with one of the following three regimens as adjuvant treatment after multidisciplinary board discussion according to the original response to chemotherapy and clinical conditions after the operation: (1) Conventional chemotherapy, (2) PD-1 blockade monotherapy, or (3) chemotherapy combined with PD-1 blockade. (Table 3)

**Table 3.** Postoperative oncological outcomes of patients receiving tumor resection.

|  | Initial N2-N3 NSCLC that had surgery<br>(n=33) |
| --- | --- |
| **Adjuvant treatment regimens with IO-no.** |  |
| Immunotherapy | 3 (9.1%) |
| Immunochemotherapy | 17 (27.3%) |
| Chemotherapy | 12 (36.4%) |
| Chemo-radiotherapy | 1 (3.0%) |
| **Median follow-up time-month (IQR)** | 9.6 (7.9–11.7) |
| **Recurrence rate at sensor-no. (%)** | 9 (27.3%) |
| **Recurrence type-no. (%)** |  |
| Bone metastasis | 2 (6.1%) |
| Brain metastasis | 2 (6.1%) |
| Lung nodule/mass | 2 (6.1%) |
| Lymph node metastasis | 3 (9.1%) |
| **Mortality at sensor-no. (%)** | 3 (9.1%) |

### Data collection, evaluation, and statistical analyses

Data were extracted independently by two investigators (HD and HL), and conflicts were adjudicated by a third investigator (WL). Information on all available variables was extracted. The following outcomes of all patients included were used to respectively assess the efficacy and safety of immunochemotherapy: (a) Radiological-regression rate and therapeutic evaluation with RECIST 1.1 standard; (b) Stage change outcomes, defined as the hilar/mediastinal/supraclavicular lymph node regression; (d) pathological regression outcomes; (e) serum tumor markers outcomes; (f) baseline details.

The tumor radiological response was evaluated through the Response Evaluation Criteria for Solid Tumors (RECIST) 1.1[13]. Calculation formula of tumor radiologic-regression rate: the longest diameter of the tumor after induction immunochemotherapy treatment, divided by the longest diameter of the tumor before immunochemotherapy treatment. The preoperative and postoperative staging was evaluated in accordance with the 8th American Joint Committee on Cancer (AJCC) lung cancer staging manuals on the tumor, node, and metastasis (TNM) staging systems[14]. Pathological analyses were performed on available biospecimens of the surgical group by two senior pathologists. MPR was defined as 10% or less viable tumor remaining on postoperative pathological review[15], while no residual tumor cells found in dissected tissues and lymph node was defined as pCR [15]. The histologic subtype was determined by a review of biopsy specimens obtained before immunochemotherapy for patients with no viable residual tumor.

Continuous data are presented as mean and standard deviation and were analyzed with 2-sample Student t-tests for independent data. Categorical variables are given as a count and percentage of patients and compared with the X^2^ or Fisher exact test. All tests were 2-sided, with an a-level of 0.05. SPSS software (SPSS version 25.0; IBM Corp, Armonk, NY) was used for all statistical evaluations. Pearson chi-square or Fisher’s exact test were used to comparing proportions. Reported P values are two-sided, and the significance level was set at 0.05 for all analyses unless otherwise noted.

## Results

### Patients’ characteristic

From January 2017 through October 2019, 55 patients with initial N2-3 metastatic NSCLC (16 patients with stage IIIA NSCLC, 30 with stage IIIB NSCLC and 9 with stage IIIC NSCLC) were eligible for inclusion in the study. Detailed baseline characteristics of the included patients were summarized in Table 1. Informed consent of patients was waived considering the retrospective setting.

**Table 1.**
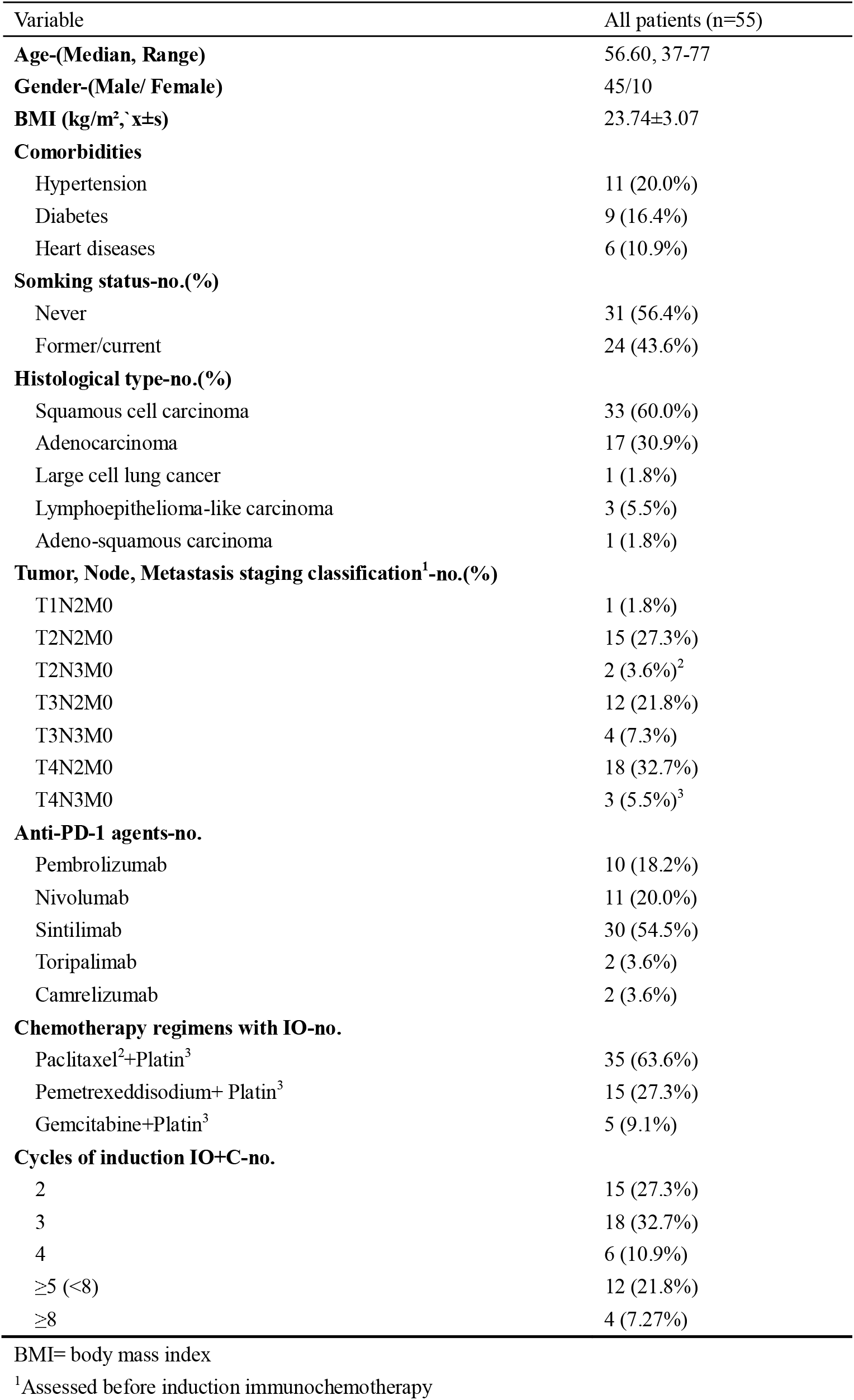
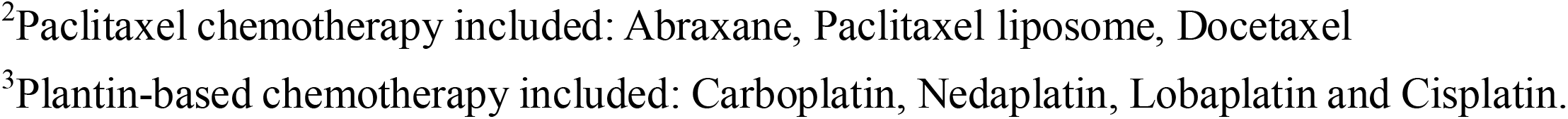
Baseline Characteristics of patients with initial stage III (N2, N3) NSCLC.

| Variable | All patients (n=55) |
| --- | --- |
| **Age-(Median, Range)** | 56.60, 37-77 |
| **Gender-(Male/ Female)** | 45/10 |
| **BMI (kg/m<sup>2</sup>, $\bar{x}\pm s$)** | 23.74±3.07 |
| **Comorbidities** |  |
| Hypertension | 11 (20.0%) |
| Diabetes | 9 (16.4%) |
| Heart diseases | 6 (10.9%) |
| **Smoking status-no.(%)** |  |
| Never | 31 (56.4%) |
| Former/current | 24 (43.6%) |
| **Histological type-no.(%)** |  |
| Squamous cell carcinoma | 33 (60.0%) |
| Adenocarcinoma | 17 (30.9%) |
| Large cell lung cancer | 1 (1.8%) |
| Lymphoepithelioma-like carcinoma | 3 (5.5%) |
| Adeno-squamous carcinoma | 1 (1.8%) |
| **Tumor, Node, Metastasis staging classification<sup>1</sup>-no.(%)** |  |
| T1N2M0 | 1 (1.8%) |
| T2N2M0 | 15 (27.3%) |
| T2N3M0 | 2 (3.6%)<sup>2</sup> |
| T3N2M0 | 12 (21.8%) |
| T3N3M0 | 4 (7.3%) |
| T4N2M0 | 18 (32.7%) |
| T4N3M0 | 3 (5.5%)<sup>3</sup> |
| **Anti-PD-1 agents-no.** |  |
| Pembrolizumab | 10 (18.2%) |
| Nivolumab | 11 (20.0%) |
| Sintilimab | 30 (54.5%) |
| Toripalimab | 2 (3.6%) |
| Camrelizumab | 2 (3.6%) |
| **Chemotherapy regimens with IO-no.** |  |
| Paclitaxel<sup>2</sup>+Platin<sup>3</sup> | 35 (63.6%) |
| Pemetrexedisodium+ Platin<sup>3</sup> | 15 (27.3%) |
| Gemcitabine+Platin<sup>3</sup> | 5 (9.1%) |
| **Cycles of induction IO+C-no.** |  |
| 2 | 15 (27.3%) |
| 3 | 18 (32.7%) |
| 4 | 6 (10.9%) |
| $\geq 5$ (<8) | 12 (21.8%) |
| $\geq 8$ | 4 (7.27%) |

BMI= body mass index
<sup>1</sup>Assessed before induction immunochemotherapy
<sup>2</sup>Paclitaxel chemotherapy included: Abraxane, Paclitaxel liposome, Docetaxel
<sup>3</sup>Platin-based chemotherapy included: Carboplatin, Nedaplatin, Lobaplatin and Cisplatin.

### Tumor and Nodal downstage efficacy of induction immunochemotherapy

For 55 patients with initial stage III (N2-3 metastatic) NSCLC, after induction immunochemotherapy, 25/55 (45.5%) patients had a partial response (PR), 23/55 (41.8%) had stable disease (SD), while 4/39 (7.3%) had PD. (Figure 2A, 2B).

**Figure 2.**
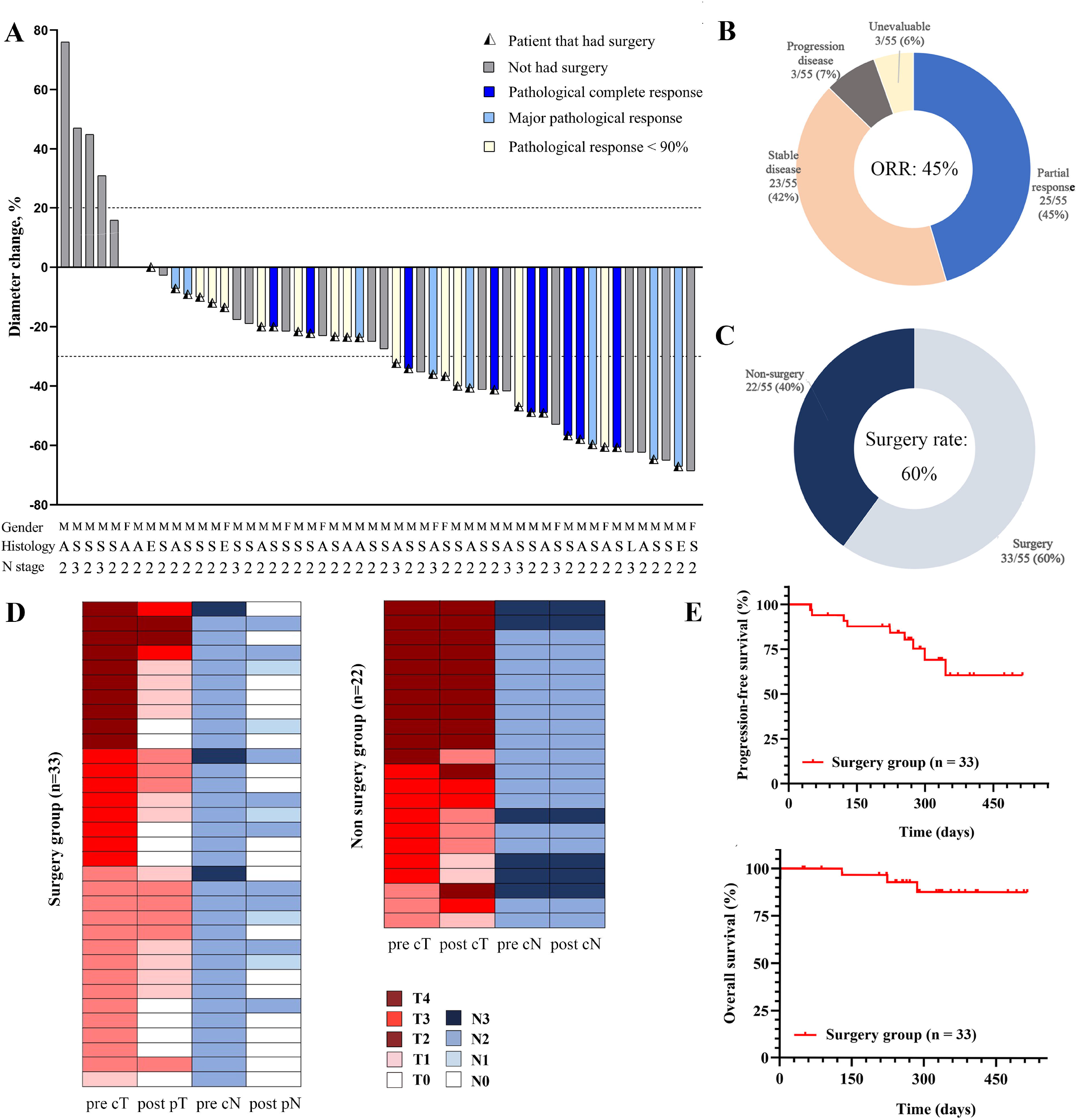
Tumor diameter change, pathological outcomes and postoperative survival outcomes of enrolled patients. (A) Tumor diameter change (%) during curative-intent induction immunochemotherapy by subgroups in 55 patients with initial unresectable N2-3 NSCLC, with each bar represents one patient. The 3 rows below the x axis shows clinical characteristics and initial lymph node(s) stage. After induction immunochemotherapy, 25/55 (45.5%) patients had partial response (PR), 23/55 (41.8%) had stable disease (SD), while 4/39 (7.3%) had PD. Of 33 patients with initial unresectable N2-3 NSCLC that underwent surgery, MPR occurred in 18 of 33 (54.5%) resected tumors, of which 10 (30.3%) specimens were considered pCR; M, male; F, female; A, lung adenocarcinoma; S, lung squamous-carcinoma; E, lung lymphoepitheliomatoid carcinoma; L, large cell lung cancer; 2, initial N2 stage; 3, initial N3 stage. (B up) Pie graph showing the oncological outcomes and (B down) percentage of patients that received surgery. (C left) Clinical stage of patient before and postoperative pathological stage after induction immunochemotherapy of patients in surgery group. (C right) Clinical stage of patient before and after immunochemotherapy of patients in nonsurgery group (D) Kaplan-Meier curves of progression-free survival and (E) overall survival in the patients included (n=33).

For 33 of 55 (60.0%) patients that underwent tumor resection (Figure 2C) after immunochemotherapy, pathological-confirmed T stage downstaging occurred in 26 (78.8%) of resected NSCLC, and 24 (72.7%) of patients had pathological-confirmed lymph node downstaging (N2-3 to N0-1). (Figure 2D left) For nine patients with initial N3 NSCLC, only 3 (33.3%) had pathological confirmed supraclavicular lymph node downstaging after immunochemotherapy and underwent surgery. Detailed induction immunochemotherapy regimens and pathologic downstaging of patients that received pulmonary resection were listed in Supplementary Table 1.

Twenty-two patients with initial N2-3 NSCLC did not undergo surgery after induction immunochemotherapy, with only 7 of whom had cT stage downstaged and no patients had cN stage downstaged. (Figure 2D right). Detailed oncological outcomes and reasons for not underwent planned surgery were summarized in Supplementary Table 3.

### Relationship between immunochemotherapy cycles and pathological response

A binary logistics analysis for MPR including the factors as follow: (1) Histological type; (2) Clinical T stage; (3) Clinical N stage; (4) Induction immunochemotherapy cycles; (Figure 3). The results showed that patients who underwent 3-4 cycles of induction immunochemotherapy were more likely to get MPR compared with conventional two cycles [Exp (B) (95% CI): 14.06 (1.59-124.08); p = 0.017]. However, adding more cycles (≥5 cycles) did not prone to better pathological response (p=0.845).

**Figure 3.**
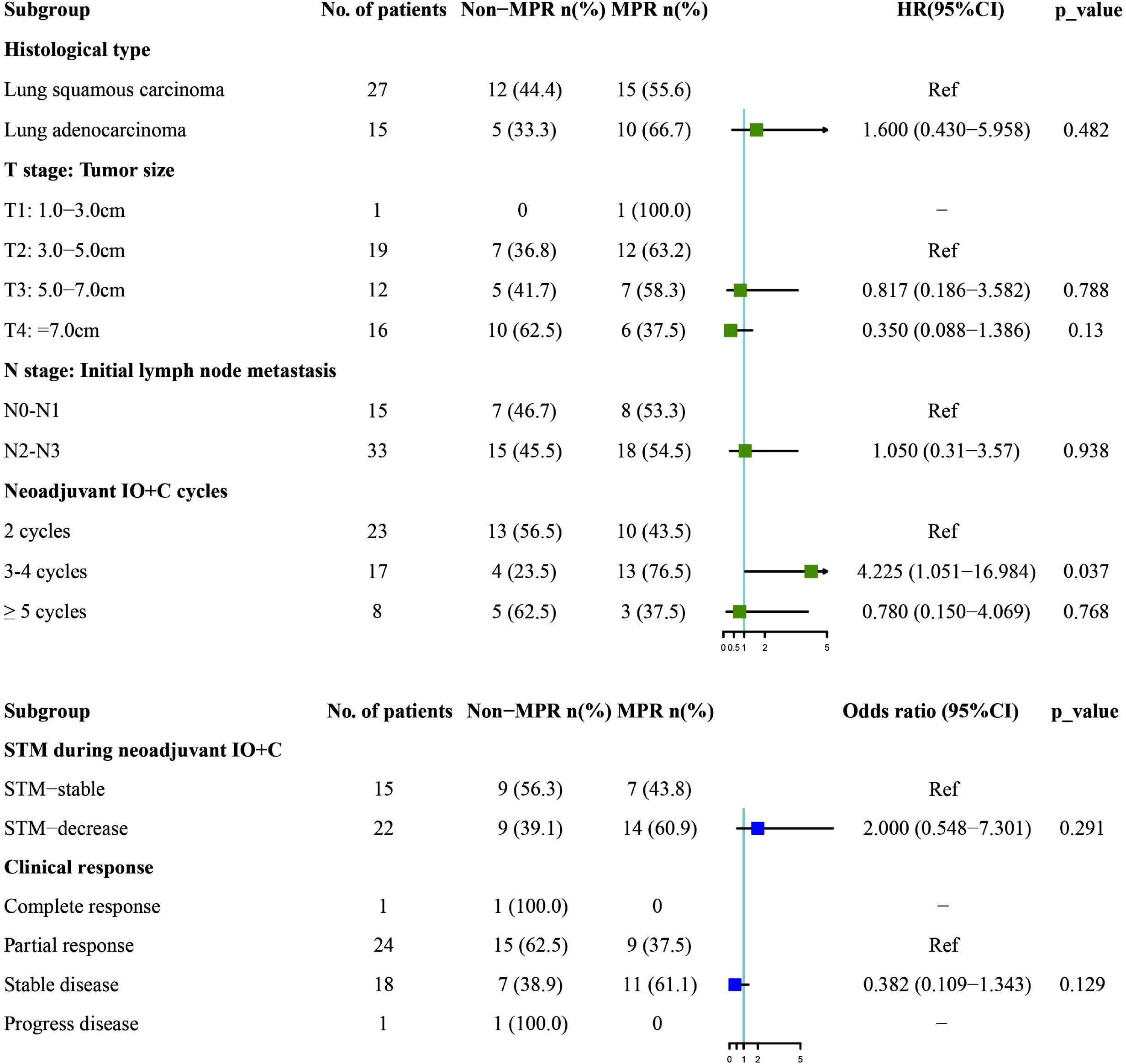
Clinicopathological characteristics and univariable analysis for major pathological response of patients that had surgery. Patients with serum tumor markers (STM): CEA, CA125, or CA153 decreased≥20% than the baseline during the induction IO+C were considered as “ STM-decrease”, or else “ STM-stable” Four patients were associated with atelectasis or obstructive pneumonitis that extends to the hilar region, making it unevaluable for the clinical response, thus they were not counted.

### Relationship between clinical response, STM, and pathological response

A univariable analysis for MPR, including the factors (1) STM change during induction immunochemotherapy; (2) Clinical response, was conducted. (Figure 3) Serum tumor markers (STM): CEA, CA125, or CA153 decreased≥20% than the baseline during the induction immunochemotherapy (p=0.291), and partial response after induction immunochemotherapy were associated with MPR (p=0.129); however, a significant difference was not reached.

### Antitumor response heterogeneity existed between LN and primary lesion

MPR occurred in 18 of 33 (54.5%) resected tumors, of which 10 (30.3%) specimens were considered pCR. Antitumor response heterogeneity existed between lymph nodes and primary tumors. Table 2 showed the clinical response and lymph node downstage status. Of 18 patients considered MPR, 4 (22.2%) patients had no lymph node downstage. While for 19 patients that had lymph node downstage (resected lymph node had no residual tumor cells, 7 (36.8%) patients still had residual tumor > 10% in primary lesion.

**Table 2.** Clinical response and lymph node (LN) downstage status in 33 patients with initial N2-3

| Variable* | No LN<br>downstage<br>(n=9) | LN downstage to<br>N1<br>(n=5) | LN downstage to<br>N0<br>(n=19) |
| --- | --- | --- | --- |
| **Major pathologic response** | 4 (44.4) | 2 (40.0) | 12 (63.2) |
| Complete pathologic response | 2 (22.2) | 1 (20.0) | 7 (36.8) |
| **Residual tumor > 10%** | 5 (55.5) | 3 (60.0) | 7 (36.8) |

We present 2 cases with typical antitumor response heterogeneity. (1) Patient 45 was diagnosed with lung squamous cell carcinoma (TNM stage: T3N2M0, stage IIIB) and received 2cycles of induction Pembrolizumab+Paclitaxel liposome+Nedaplatin. The patient then underwent lobectomy of RUL, and the resected specimen achieved 100% pathologic remission (Figure 4A left) but was identified hardly any pathologic remission at resected 4R lymph node (Figure 4A left). (2) Patient 52 was diagnosed with lung squamous cell carcinoma (TNM stage: T4N2M0, stage IIIB) and received 2cycles of induction Sintilimab+Abraxane+Carboplatin. The patient then underwent lobectomy of RUL, and the resected specimen 2R lymph node showed no residual tumor cell (Figure 4B right). However, in the resected specimen primary tumor, only 20% pathologic remission was achieved (Figure 4B left).

**Figure 4.**
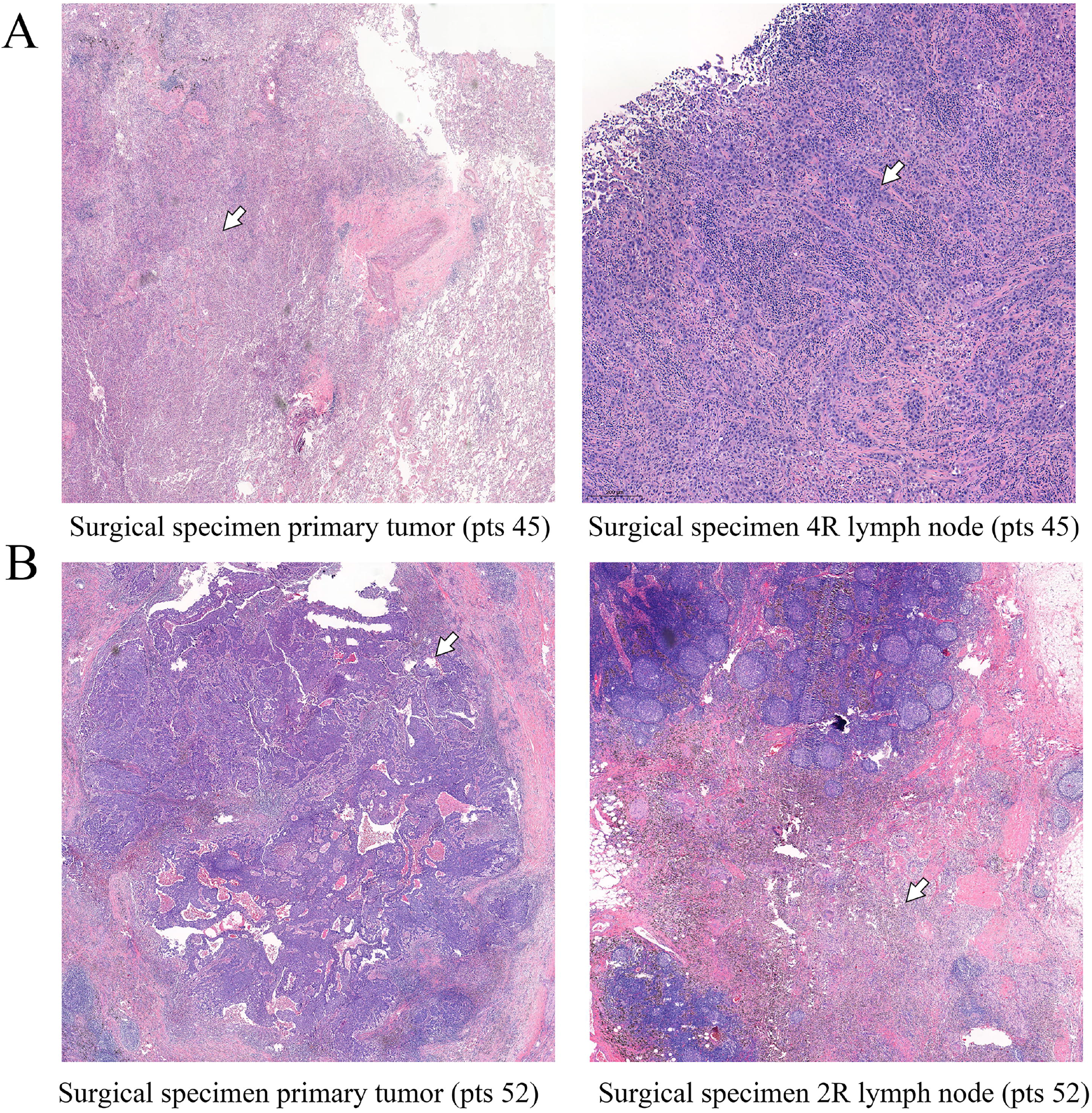
Special cases report demonstrating different response between primary tumor and metastatic lymph node. (A) A patient with age ranges from 55-60 was diagnosed as lung squamous cell carcinoma (TNM stage: T3N2M0, stage IIIB) and received 2cycles of induction Pembrolizumab+Paclitaxel liposome+Nedaplatin. The patient then underwent lobectomy of RUL. The pathologic images shown (A-left) are the primary tumor and large amount of inflammatory cell was found, with 100% pathologic remission been achieved (white arrow); (A-right) hardly no pathologic remission at resected 4R lymph node (white arrow). (B) A patient with age ranges from 55-60 diagnosed as lung squamous cell carcinoma (TNM stage: T4N2M0, stage IIIB) and received 2cycles of induction Sintilimab+Abraxane+Carboplatin. The patient then underwent lobectomy of RUL. The pathologic images shown (B-left) are the primary tumor specimen with 80% residual tumor cell left (white arrow). While the (B-right) resected specimen 2R lymph node showed no residual tumor cell (white arrow).

### Survival outcomes

The Kaplan–Meier curves for progression-free survival (Figure 2E) and overall survival (Figure 2D) in the study population that had surgery were shown in Figure 3. Table 4 showed the postoperative oncological outcomes of 48 patients receiving tumor resection. At the time of data cutoff (October 30th, 2020), the median follow-up duration was 9.6 months (IQR 7.9–11.7), 24 of 33 (72.7%) patients that had pulmonary resection were progression-free. Thirty of them (90.9%) still alive at the time cutoff, while two patients died of lymph node relapse 223 days and 129 days after surgery, respectively, and one patient died of brain metastasis 255 days after surgery (Supplementary Table 3). The estimated 1-year progression-free survival was 60.14% and the 1-year overall survival was 87.78%. (Table 3)

## Discussion

Through immunochemotherapy, 25/55 (45.5%) patients with stage III (N2, N3) NSCLC in the current study had PR. With the aim of increasing both locoregional and systemic control, radical tumor resections were administrated after induction immunochemotherapy in selected 33 patients. Respecting efficacy, MPR occurred in 18 of 33 (54.5%) resected tumors and pCR in 10 (30.3%) patients. Compared with the SAKK16/14 study [19], a slightly lower MPR rate (54.5% versus 60%), ORR (45.5% versus 58.1%) and similar 12-month EFS (72.7% vs 71.3%) were shown in this study. However, the calculated MPR rate (54.5% vs. 85.4%) and the estimate PFS (72.7% vs. 95.7%) at 12 month was worse than that of the NADIM[20] trial, this might be due to the fact that most of the patients included in this study were with IIIB NSCLC, and potential micrometastasis tends to be accompanied with more advanced stage tumor. To be noted, 16 patients did not undergo surgery, with a majority of them had no tumor downstaging or with tumor progression; thus, the actual response rate of all 55 patients with initial N2-N3 lymph node would be even lower.

This study also showed that the antitumor activity could be different between metastatic lymph node and primary tumor in patients with unresectable N2/3 NSCLC: 4 patients had MPR of the primary lesion but with no lymph node downstage, while seven patients had no metastatic tumor cells in resected lymph nodes but with pathological response<90% in primary lesion. Gao et al.[17] administrated 2cycles of induction sintilimab in resectable NSCLC and identified different responses between primary tumors and lymph node metastases in 18 patients. Liu et al. [18] reported that induction PD-1 blockade could enhance the systemic priming of antitumor T cells to eradicate distant metastases. However, whether primed T cell might be impeded from infiltrating into the lymph nodes or any up-regulated molecular markers affecting the unmatched response remains unknown. The immune microenvironment between primary cancer and metastatic lymph nodes might also lead to this phenomenon, and further study could focus on how to reverse the “ local resistance.”

Notably, a patient (pts 71) in this study experienced progress disease (PD) according to RECIST 1.1 after two cycles of induction immunochemotherapy. However, this patient still had complete pathologic remission (pCR). This may be related to massive fibrosis, lymphocytic infiltration, and peritumoral inflammation occupying the original tumor location after tumor retraction instead of tumor growth. This pseudo-progression was also reported by Tanizaki et al. and Bott et al. [21, 22], in which two pCR patients only had stable disease during treatment of induction IO. The result demonstrated that a proportion of patients with locally advanced NSCLC could benefit from induction IO without initial radiographic tumor shrinkage, even it presented as PD. A more comprehensive response evaluation of induction IO combining CT, SUV value, and serum tumor markers could be useful for identifying this phenomenon before surgery.

Considering that some patients have received more than four courses, including ≥5-8 cycles (∼30% of N2-N3 patients) in this study. We conducted a univariable analysis to investigate the association of clinical factors with MPR. And the result demonstrated that patients who underwent 3-4 cycles of induction immunochemotherapy were more likely to get MPR compared with the conventional two cycles (p = 0.017). In clinical trials regarding induction IO in early-stage NSCLC [23, 24], two cycles of IO were administrated before surgery. However, a recent article demonstrated that: a proportion of the top 1% of intra-tumor clonotypes shared with the peripheral T cell receptor repertoire significantly increased after the second cycle of the preoperative anti-PD-1 agent, and the upward trend side remained. The results indicated that the antitumor response is still growing at the second cycle of preoperative PD-1 blockade[25], which reinforced the necessity to extend the induction immunochemotherapy cycle for achieving MPR.

Nevertheless, we also found that adding more cycles based on four cycles (≥ 5 cycles) was not associating with the presence of MPR (p=0.845). In our center, a strategy of offering more cycles until the tumor was down-staged and became resectable was preferred. And it indicated that some patients might still not respond to induction immunochemotherapy even though sufficient time and cycles were given.

In this study, of 33 patients who underwent surgery, 15 (45.5%) reported different levels of pleural adhesion, which can be regarded as a post-immunotherapy response. Chaft et al. [10] included five patients with advanced NSCLC that underwent pneumonectomy after treatment with t-cell checkpoint inhibitors; mediastinal and hilar fibrosis could be seen intraoperatively. Bott et al. [21] investigated pneumonectomy after induction immunotherapy in resectable NSCLC, and more than half of the VATS cases were with perihilar inflammation and fibrosis. Furthermore, the brittleness of the vessels did contribute to the increased difficulty of the operation.

Several limitations of this study should be acknowledged. Firstly, this study did not investigate immune-related adverse events of the immunochemotherapy, which influences the tolerance of induction immunochemotherapy. Secondly, we stage each patient before the operation with PET plus contrast-enhanced CT (few of them had EBUS), which might not as accurate as mediastinoscopy; Thirdly, patients in the nonsurgery group could not be staged through surgical specimens, which could bring bias to the integrated downstaging rate considering the pseudo-progression phenomenon and the false positive rate of PET/CT scan stage for lymph nodes.

## Conclusions

Immunochemotherapy in patients with stage III (N2, N3) NSCLC is feasible for tumor and nodal downstaging. We believe that the indications of induction immunochemotherapy can be further expanded to initial stage III NSCLC in strictly selected patients, given the acceptable recurrence risk and surgery-related mortality shown above. For initial N2/3 NSCLC, the antitumor response could differ between metastatic nodals and primary tumors. Prolonged cycles of immunochemotherapy (3-4 cycles) were more appropriate for stage III (N2, N3) NSCLC than 1-2 cycles for higher tumor radiologic-regression rate and MPR rate.

## Data Availability

All data included in this study are available upon request by contact with the corresponding author.

## References

1. Bradley JD, Paulus R, Komaki R et al. Standard-dose versus high-dose conformal radiotherapy with concurrent and consolidation carboplatin plus paclitaxel with or without cetuximab for patients with stage IIIA or IIIB non-small-cell lung cancer (RTOG 0617): a randomised, two-by-two factorial phase 3 study. Lancet Oncol 2015; 16: 187–199.

2. Shu CA, Gainor JF, Awad MM et al. Neoadjuvant atezolizumab and chemotherapy in patients with resectable non-small-cell lung cancer: an open-label, multicentre, single-arm, phase 2 trial. Lancet Oncol 2020; 21: 786–795.

3. Gaudreau PO, Negrao MV, Mitchell KG et al. Neoadjuvant Chemotherapy Increases Cytotoxic T Cell, Tissue Resident Memory T Cell, and B Cell Infiltration in Resectable NSCLC. J Thorac Oncol 2021; 16: 127–139.

4. Tfayli A, Al Assaad M, Fakhri G et al. Neoadjuvant chemotherapy and Avelumab in early stage resectable non-small cell lung cancer. Cancer Med 2020; 9: 8406–8411.

5. Reuss JE, Anagnostou V, Cottrell TR et al. Neoadjuvant nivolumab plus ipilimumab in resectable non-small cell lung cancer. J Immunother Cancer 2020; 8.

6. Provencio M, Nadal E, Insa A et al. Neoadjuvant chemotherapy and nivolumab in resectable non-small-cell lung cancer (NADIM): an open-label, multicentre, single-arm, phase 2 trial. Lancet Oncol 2020; 21: 1413–1422.

7. Gettinger S, Horn L, Jackman D et al. Five-Year Follow-Up of Nivolumab in Previously Treated Advanced Non-Small-Cell Lung Cancer: Results From the CA209-003 Study. J Clin Oncol 2018; 36: 1675–1684.

8. Horn L, Spigel DR, Vokes EE et al. Nivolumab Versus Docetaxel in Previously Treated Patients With Advanced Non-Small-Cell Lung Cancer: Two-Year Outcomes From Two Randomized, Open-Label, Phase III Trials (CheckMate 017 and CheckMate 057). J Clin Oncol 2017; 35: 3924–3933.

9. Gray JE, Villegas A, Daniel D et al. Brief report: Three-year overall survival with durvalumab after chemoradiotherapy in Stage III NSCLC - Update from PACIFIC. J Thorac Oncol 2019.

10. Chaft JE, Hellmann MD, Velez MJ et al. Initial Experience With Lung Cancer Resection After Treatment With T-Cell Checkpoint Inhibitors. Ann Thorac Surg 2017; 104: e217–e218.

11. Liang H, Yang C, Gonzalez-Rivas D et al. Sleeve lobectomy after neoadjuvant chemoimmunotherapy/chemotherapy for local advanced non-small cell lung cancer. Transl Lung Cancer Res 2021; 10: 143–155.

12. Ettinger DS, Wood DE, Aggarwal C et al. NCCN Guidelines Insights: Non-Small Cell Lung Cancer, Version 1.2020. J Natl Compr Canc Netw 2019; 17: 1464–1472.

13. Eisenhauer EA, Therasse P, Bogaerts J et al. New response evaluation criteria in solid tumours: revised RECIST guideline (version 1.1). Eur J Cancer 2009; 45: 228–247.

14. Goldstraw P, Chansky K, Crowley J et al. The IASLC Lung Cancer Staging Project: Proposals for Revision of the TNM Stage Groupings in the Forthcoming (Eighth) Edition of the TNM Classification for Lung Cancer. Journal of Thoracic Oncology 2016; 11: 39–51.

15. Pataer A, Kalhor N, Correa AM et al. Histopathologic response criteria predict survival of patients with resected lung cancer after neoadjuvant chemotherapy. J Thorac Oncol 2012; 7: 825–832.

16. Aupérin A, Le Péchoux C, Rolland E et al. Meta-analysis of concomitant versus sequential radiochemotherapy in locally advanced non-small-cell lung cancer. J Clin Oncol 2010; 28: 2181–2190.

17. Gao S, Li N, Gao S et al. Neoadjuvant PD-1 inhibitor (Sintilimab) in NSCLC. Journal of Thoracic Oncology 2020; 15: 816–826.

18. Liu J, Blake SJ, Yong MC et al. Improved Efficacy of Neoadjuvant Compared to Adjuvant Immunotherapy to Eradicate Metastatic Disease. Cancer Discov 2016; 6: 1382–1399.

19. Rothschild S, Zippelius A, Savic S et al. SAKK 16/14: Anti-PD-L1 antibody durvalumab (MEDI4736) in addition to neoadjuvant chemotherapy in patients with stage IIIA(N2) non-small cell lung cancer (NSCLC)—A multicenter single-arm phase II trial. Journal of Clinical Oncology 2018; 36: TPS8584–TPS8584.

20. Provencio M, Nadal E, Insa A et al. OA01.05 Phase II Study of Neo-Adjuvant Chemo/Immunotherapy for Resectable Stages IIIA Non-Small Cell Lung Cancer-Nadim Study-SLCG. Journal of Thoracic Oncology 2018; 13: S320.

21. Bott MJ, Yang SC, Park BJ et al. Initial results of pulmonary resection after neoadjuvant nivolumab in patients with resectable non-small cell lung cancer. J Thorac Cardiovasc Surg 2019; 158: 269–276.

22. Tanizaki J, Hayashi H, Kimura M et al. Report of two cases of pseudoprogression in patients with non-small cell lung cancer treated with nivolumab-including histological analysis of one case after tumor regression. Lung Cancer 2016; 102: 44–48.

23. Forde PM, Chaft JE, Smith KN et al. Neoadjuvant PD-1 Blockade in Resectable Lung Cancer. N Engl J Med 2018; 378: 1976–1986.

24. Kwiatkowski DJ, Rusch VW, Chaft JE et al. Neoadjuvant atezolizumab in resectable non-small cell lung cancer (NSCLC): Interim analysis and biomarker data from a multicenter study (LCMC3). Journal of Clinical Oncology 2019; 37: 8503–8503.

25. Zhang J, Ji Z, Caushi JX et al. Compartmental analysis of T cell clonal dynamics as a function of pathologic response to neoadjuvant PD-1 blockade in resectable non-small cell lung cancer. Clin Cancer Res 2019.

