## Supplementary Table 1 for "Immunochemotherapy as induction treatment in Stage III (N2, N3) Non-small cell lung cancer"

**Supplementary Table 1**. Detailed neoadjuvant IO+C regiments and pathologic downstaging of patients that received pulmonary resection

| Patients number | Histology | PD-1 inhibitor | Chemotherapy regiments | cTNM | Cycles | Clinical response | IO+C duration^1^ (d) | pTNM | Stage | Pathologic response of primary lesion | Lymph node downstage |
| --- | --- | --- | --- | --- | --- | --- | --- | --- | --- | --- | --- |
| 1 | LUSQ | Pembrolizumab | Paclitaxel liposome+Carboplatin | T3N2M0 | 3 | PR | 81 | T1N2M0 | IIIA | Major response | N0 |
| 2 | LUAD | Pembrolizumab | Pemetrexeddisodium+Carboplatin | T3N3M0 | 3 | PR | 69 | T2N2M0 | IIIA | <90% | N0 |
| 10 | LUAD | Sintilimab | Pemetrexeddisodium+Carboplatin | T2N3M0 | 5 | PR | 207 | T1N0M0 | IA | Major response | Yes |
| 11 | LUSQ | Nivolumab | Paclitaxel liposome+Carboplatin | T3N2M0 | 3 | PR | 105 | T1N1M0 | IIB | Major response | Yes |
| 19 | LELC | Sintilimab | Paclitaxel liposome+Carboplatin | T4N2M0 | 5 | SD | 131 | T4N0M0 | IIIA | <90% | Yes |
| 24 | LUAD | Pembrolizumab | Pemetrexeddisodium+Carboplatin | T4N2M0 | 2 | PR | 67 | T1N1M0 | IIB | <90% | Yes |
| 26 | LUAD | Sintilimab | Abraxane+Cisplatin | T4N2M0 | 6 | SD | 182 | T1N0M0 | IA | Major response | Yes |
| 33 | LUSQ | Sintilimab | Abraxane+Cisplatin | T4N3M0 | 10 | PR | 461 | T3N0M0 | IIB | <90% | Yes |
| 34 | LUSQ | Sintilimab | Abraxane+Carboplatin | T2N2M0 | 2 | PR | 69 | T2N0M0 | IIA | <90% | Yes |
| 36 | LUAD | Pembrolizumab | Pemetrexeddisodium+Lobaplatin | T2N2M0 | 2 | Unevaluable^2^ | 80 | T0N2M0 | IIIA | Complete response | NO |
| 38 | LUSQ | Sintilimab | Abraxane+Carboplatin | T2N2M0 | 5 | SD | 138 | T0N0M0 | - | Complete response | Yes |
| 41 | LUAD | Toripalimab | Pemetrexeddisodium+Nedaplatin | T2N2M0 | 2 | PR | 58 | T1N0M0 | IA | Major response | Yes |
| 42 | LUSQ | Camrelizumab | Paclitaxel liposome+Carboplatin | T4N2M0 | 13 | SD | 463 | T3N2M0 | IIIB | <90% | N0 |
| 44 | LUAD | Sintilimab | Pemetrexeddisodium+Lobaplatin | T1N2M0 | 3 | PR | 110 | T0N0M0 | - | Complete response | Yes |
| 45 | LUSQ | Pembrolizumab | Paclitaxel liposome+Nedaplatin | T3N2M0 | 2 | SD | 56 | T0N2M0 | IIIA | Complete response | N0 |
| 46 | LELC | Sintilimab | Gemcitabine+Nedaplatin | T4N2M0 | 3 | PR | 62 | T1N0M0 | IA | Major response | Yes |
| 47 | LUSQ | Nivolumab | Abraxane+Carboplatin | T2N2M0 | 3 | PR | 58 | T0N0M0 | - | Complete response | Yes |
| 49 | LUSQ | Nivolumab | Paclitaxel liposome+Carboplatin | T2N2M0 | 4 | SD | 121 | T1N2M0 | IIIA | Major response | N0 |
| 50 | LUAD | Nivolumab | Paclitaxel liposome+Carboplatin | T4N2M0 | 3 | Unevaluable^2^ | 112 | T4N2M0 | IIIB | <90% | N0 |
| 51 | LUSQ | Sintilimab | Abraxane+Nedaplatin | T2N2M0 | 2 | SD | 61 | T2N1M0 | IIB | <90% | Yes |
| 52 | LUSQ | Sintilimab | Abraxane+Carboplatin | T4N2M0 | 2 | PR | 68 | T0N1M0 | IIB | Complete response | Yes |
| 53 | LUSQ | Nivolumab | Paclitaxel liposome+Nedaplatin | T2N2M0 | 2 | SD | 63 | T1N1M0 | IIB | <90% | Yes |
| 56 | LELC | Pembrolizumab | Gemcitabine+Nedaplatin | T2N2M0 | 2 | SD | 76 | T2N2M0 | IIIA | <90% | N0 |
| 57 | LUAD | Sinitilumab | Pemetrexeddisodium+Lobaplatin | T3N2M0 | 2 | PR | 58 | T0N0M0 | - | Complete response | Yes |
| 58 | LUSQ | Sinitilumab | Paclitaxel liposome+Carboplatin | T3N2M0 | 2 | PR | 52 | T0N0M0 | - | Complete response | Yes |
| 59 | LASC | Nivolumab | Gemcitabine+Carboplatin | T3N2M0 | 3 | SD | 452 | T2N0M0 | IIA | <90% | Yes |
| 61 | LUSQ | Toripalimab | Paclitaxel liposome+Cisplatin | T2N2M0 | 3 | PR | 90 | T0N0M0 | - | Complete response | Yes |
| 63 | LUSQ | Sinitilumab | Paclitaxel liposome+Carboplatin | T3N2M0 | 5 | SD | 144 | T2N0M0 | IIA | <90% | Yes |
| 64 | LUSQ | Sinitilumab | Paclitaxel liposome+Lobaplatin | T4N2M0 | 3 | PR | 68 | T1N0M0 | IA | <90% | Yes |
| 65 | LUSQ | Sinitilumab | Paclitaxel liposome+Carboplatin | T4N2M0 | 3 | PR | 103 | T0N0M0 | - | Complete response | Yes |
| 68 | LUAD | Camrelizumab | Pemetrexeddisodium+Carboplatin | T2N2M0 | 3 | SD | 102 | T1N0M0 | IA | Major response | Yes |
| 69 | LUAD | Toripalimab | Paclitaxel liposome+Carboplatin | T2N2M0 | 2 | SD | 79 | T2N2M0 | IIIA | <90% | N0 |
| 70 | LUSQ | Sinitilumab | Paclitaxel liposome+Carboplatin | T2N2M0 | 2 | Unevaluable^2^ | 59 | T2N0M0 | IIA | <90% | Yes |

^1^The interval from the first cycle of induction immunochemotherapy to surgery.

^2^Four patients were associated with atelectasis or obstructive pneumonitis that extends to the hilar region, making it unevaluable for the clinical response.
