## Supplementary Table 2 for "Immunochemotherapy as induction treatment in Stage III (N2, N3) Non-small cell lung cancer"

**Supplementary Table S2.** Detailed survival time and progression-free survival time of patients that had surgery.

| Patients number | Survival (days) | Survival status | Progression-free survival (days) | Progression status |
| --- | --- | --- | --- | --- |
| 1 | 507.00 | survive | 299.00 | progressed^3^ |
| 2 | 223.00 | dead^1^ | 223.00 | progressed^l^ |
| 8 | 614.00 | survive | 614.00 | progression-free |
| 10 | 514.00 | survive | 514.00 | progression-free |
| 11 | 411.00 | survive | 345.00 | progressed^3^ |
| 12 | 568.00 | survive | 568.00 | progression-free |
| 16 | 500.00 | survive | 500.00 | progression-free |
| 19 | 290.00 | survive | 290.00 | progression-free |
| 22 | 332.00 | survive | 332.00 | progression-free |
| 23 | 360.00 | survive | 99.00 | progressed^4^ |
| 24 | 492.00 | survive | 492.00 | progression-free |
| 25 | 476.00 | survive | 476.00 | progression-free |
| 26 | 355.00 | survive | 355.00 | progression-free |
| 30 | 349.00 | survive | 349.00 | progression-free |
| 33 | 86.00 | survive | 86.00 | progression-free |
| 34 | 474.00 | survive | 474.00 | progression-free |
| 36 | 285.00 | dead^2^ | 255.00 | progressed^2^ |
| 38 | 221.00 | survive | 221.00 | progression-free |
| 41 | 407.00 | survive | 407.00 | progression-free |
| 42 | 47.00 | survive | 47.00 | progressed^5^ |
| 44 | 372.00 | survive | 372.00 | progression-free |
| 45 | 129.00 | dead^1^ | 129.00 | progressed^6^ |
| 46 | 325.00 | survive | 274.00 | progressed^6^ |
| 47 | 330.00 | survive | 330.00 | progression-free |
| 48 | 367.00 | survive | 367.00 | progression-free |
| 49 | 337.00 | survive | 337.00 | progression-free |
| 50 | 334.00 | survive | 334.00 | progression-free |
| 51 | 385.00 | survive | 398.00 | progression-free |
| 52 | 288.00 | survive | 288.00 | progression-free |
| 53 | 263.00 | survive | 263.00 | progression-free |
| 54 | 215.00 | survive | 323.00 | progression-free |
| 55 | 269.00 | survive | 269.00 | progression-free |
| 56 | 270.00 | survive | 270.00 | progression-free |
| 57 | 255.00 | survive | 255.00 | progression-free |
| 58 | 288.00 | survive | 288.00 | progression-free |
| 59 | 51.00 | survive | 51.00 | progressed^4^ |
| 60 | 295.00 | survive | 295.00 | progression-free |
| 61 | 355.00 | survive | 121.00 | progressed^6^ |
| 62 | 296.00 | survive | 49.00 | progressed^4^ |
| 63 | 240.00 | survive | 240.00 | progression-free |
| 64 | 206.00 | survive | 206.00 | progression-free |
| 65 | 254.00 | survive | 267.00 | progression-free |
| 66 | 267.00 | survive | 267.00 | progression-free |
| 67 | 288.00 | survive | 288.00 | progression-free |
| 68 | 242.00 | survive | 242.00 | progression-free |
| 69 | 222.00 | survive | 222.00 | progression-free |
| 70 | 256.00 | survive | 256.00 | progression-free |
| 71 | 237.00 | survive | 237.00 | progression-free |

^1^Patients died of lymph node relapse.

^2^Patients died of brain metastasis.

^3^Lung nodule/mass relapsed in the lung

^4^Bone metastasis

^5^Brain metastasis

^6^Lymph node metastasis (enlarged swollen stage N1 or N2 lymph node)
