## Supplementary Table 3 for "Immunochemotherapy as induction treatment in Stage III (N2, N3) Non-small cell lung cancer"

**Supplementary Table S3**. Detailed oncological outcomes for 23 patients not underwent planned surgery.

| Patients number | Histology | cTNM | Stage | Cycle | IO+C duration (days) | Tumor size before IO+C | Tumor size after IO+C | Tumor regression rate (%)^1^ | Clinical response | Stage change |
| --- | --- | --- | --- | --- | --- | --- | --- | --- | --- | --- |
| 3 | LUAD | T3N2M0 | IIIB | 6 | 145 | 5 | 8.8 | 76 | PD | Systemic multiple metastasis |
| 4 | LUSQ | T4N2M0 | IIIC | 3 | 74 | 12.5 | 11.6 | -19 | SD | - |
| 5 | LUSQ | T3N2M0 | IIIA | 4 | 127 | 4 | 2.9 | -27.5 | SD | - |
| 6 | LUSQ | T2N2M0 | IIIB | 3 | 45 | 4.9 | 7.1 | 44.9 | PD | Tumor progressed |
| 7 | LUSQ | T4N3M0 | IIIB | 8 | 30 | 1.9 | 2.8 | 47 | PD | Tumor progressed |
| 9 | LUAD | T4N2M0 | IIIB | 2 | 190 | 6.5 | 6.5 | 0 | SD | - |
| 13 | LUSQ | T3N2M0 | IIIB | 6 | 117 | 6.3 | 2.2 | -65 | PR | T stage Downstage^2^ |
| 14 | LUSQ | T4N2M0 | IIIB | 2 | 184 | 10.8 | 3.4 | -68.52 | PR | T stage Downstage^2^ |
| 15 | LUSQ | T4N3M0 | IIIC | 9 | 199 | 6.8 | 3.2 | -52.94 | PR | - |
| 17 | LUAD | T4N2M0 | IIIB | 7 | 20 | 4.5 | 4.5 | 0 | SD | - |
| 18 | LUSQ | T4N2M0 | IIIB | 7 | 48 | 8.2 | 9.5 | 15.9 | SD | - |
| 20 | LUAD | T4N2M0 | IIIB | 3 | 147 | 6.1 | 2.3 | -62.3 | PR | T stage Downstage^3^ |
| 21 | LUAD | T3N3M0 | IIIB | 4 | 120 | 2.4 | 1.4 | -41.67 | PR | T stage Downstage^3^ |
| 27 | LUSQ | T3N2M0 | IIIB | 3 | 72 | 7.6 | 5.7 | -25 | SD | - |
| 28 | LUSQ | T4N2M0 | IIIB | 3 | 81 | 3.7 | 3.6 | -2.7 | SD | - |
| 29 | LCLC | T3N3M0 | IIIB | 5 | 156 | 5.3 | 2 | -62.26 | PR | Systemic multiple metastasis |
| 31 | LUAD | T4N2M0 | IIIC | 5 | 86 | 5.2 | 4 | -23.07 | SD | - |
| 32 | LUSQ | T3N2M0 | IIIB | 4 | 44 | 5.1 | 3 | -41.18 | PR | T stage Downstage^3^ |
| 35 | LUSQ | T2N3M0 | IIIB | 4 | 56 | 8.5 | 7 | -17.65 | SD | - |
| 37 | LUSQ | T4N2M0 | IIIC | 4 | 64 | 5.1 | 4 | -21.57 | SD | - |
| 39 | LUSQ | T3N3M0 | IIIB | 3 | 64 | 3.5 | 4.6 | 31 | PD | Tumor progressed |
| 40 | LUSQ | T2N2M0 | IIIA | 2 | 91 | 3.4 | 2.2 | -35.29 | PR | T stage Downstage^3^ |

^1^Calculation formula of tumor radiologic-regression rate: the longest diameter of the tumor after immunochemotherapy, divided by the longest diameter of the tumor before immunochemotherapy.

^2^Patients refused the surgery
